# Automatic Detection and Assessment of Freezing of Gait Manifestations

**DOI:** 10.1101/2023.07.10.23292437

**Authors:** Po-Kai Yang, Benjamin Filtjens, Pieter Ginis, Maaike Goris, Alice Nieuwboer, Moran Gilat, Peter Slaets, Bart Vanrumste

## Abstract

Freezing of gait (FOG) is an episodic and highly disabling symptom of Parkinson’s disease (PD). Although described as a single phenomenon, FOG is not univocal and can express as different manifestations, such as trembling in place or complete akinesia. We aimed to analyze the utility of deep learning trained on inertial measurement unit data to classify FOG into both manifestations. We developed a temporal convolutional neural network, which we compared to three state-of-the-art FOG detection algorithms that were adapted to the FOG manifestation detection task. Next, we investigated its performance in distinguishing between the two manifestations and other forms of movement cessation (e.g., volitional stopping and sitting) based on gold-standard video annotations. Experiments were conducted on a dataset of twelve PD patients with FOG that completed a FOG-provoking protocol, including the timed-up-and-go and 360-degree turning-in-place tasks during ON and OFF anti-Parkinsonian medication. The results showed that our model enables accurate detection of FOG manifestations with an 11.43% higher F1 score than the second-best model. Assessment of FOG manifestation severity was moderately strong for trembling in place (Intra-class Correlation Coefficient (ICC)=0.64, [0.16,0.88]) and strong for complete akinesia (ICC=0.87, [0.63,0.96]). Remarkably, our results show that complete akinesia can be distinguished from volitional stopping. In conclusion, we established that FOG manifestations could be accurately detected and assessed with deep learning. Future work should establish whether these results hold firm for a more extensive and varied verification cohort.

## I. Introduction

Parkinson’s disease (PD) is a neurodegenerative disorder that already affects over six million people worldwide with a prevalence that is rising [1]. One of the most debilitating symptoms associated with PD is freezing of gait (FOG), which has been defined as a “brief, episodic absence or marked reduction of forward progression of the feet despite the intention to walk” [1]–[3]. The unpredictable nature and the inability of patients to take corrective steps after losing their balance during FOG poses a significant risk of falls and related injuries for PD patients [4]–[6], and a lower quality of life [7]. Although described as a single phenomenon, FOG is not univocal and can be expressed as different manifestations, namely: 1) episodic rapid shuffling with very short steps and poor clearance of the feet, 2) trembling in place visible as alternating tremulous oscillations in the legs with minimal or no forward progression, and 3) complete akinesia with minimal or no visible movement in the lower limbs [8]. However, whether or not shuffling should be included in the definition of FOG is being debated given that there is still forward progression of the feet [9]. As the etiology of the different manifestations likely differs and, as such, may respond differently to therapy, developing an objective assessment of the FOG manifestations will improve our understanding of this complex symptom and help guide appropriate treatment [8].

The current study is the first attempt to automatically quantify different FOG manifestations using deep learning (DL) and lower limb movement characteristics measured by inertial measurement units (IMUs). We adjusted three state-of-the-art FOG detection algorithms to the FOG manifestations detection task. These algorithms served as a baseline for comparison with our previously validated FOG manifestation detection algorithm that was not specifically trained to detect manifestations [9]. To quantify FOG manifestation severity, we calculated the percentage time frozen (%TF) as per previous work [10], [11] and the percentage time frozen of each manifestation. Given the lack of overt movement in the legs during particularly akinetic FOG episodes, it is important to verify that the model is able to distinguish such FOG events from volitional stopping. As such, to determine the robustness of our approach, we further investigated whether our DL algorithm could distinguish between FOG manifestations and other forms of movement cessation (e.g., volitional stopping and sitting) [12].

## II. Related work

Various methods have been proposed to automatically detect and assess FOG using wearable sensor data obtained through IMUs [9], [13]–[18]. IMUs could record the movement of the associated body segment as a time series of 3-axis acceleration and angular velocity. The raw signals themselves or features extracted from them have been employed to train various FOG detection models. Based on the data segmentation method, FOG detection using IMU data can be divided into two distinct approaches: window-based and sample-based. The former uses a sliding window (usually 1 second) to segment the sensor data and extract features, while the latter detects FOG at the recorded sample level (e.g., 500 Hz).

### A. Window-based methods

Window-based methods tackle automated FOG detection as an action recognition problem [13]–[18]. These methods segment an IMU sequence into fixed-length windows using a sliding-window scheme. Within each window, a single label is predicted for all the samples as either FOG or non-FOG. Since each window can contain multiple labels at FOG and non-FOG transitions, the ground-truth label is typically established through majority voting [14]–[16].

Such earlier approaches also relied on manual feature engineering to distinguish between FOG and non-FOG. For instance, Moore et al. developed a thresholding algorithm based on the Freeze Index (FI) to distinguish between FOG and non-FOG [19]. They defined the FI as the power in the freezing band (0.5-3 Hz) divided by the power in the locomotor band (3-8 Hz), which others have subsequently applied as well [13]. However, other studies, such as Bächlin et al. and Delval et al., introduced an energy threshold and stride features, which were combined with the FI to identify FOG episodes [20], [21].

Going beyond the aforementioned threshold-based methods, previous studies also employed traditional machine learning models on hand-engineered features to detect FOG. For example, Tsipouras et al. employed decision trees and random forests on the mean entropy calculated from the acceleration of six IMUs (i.e., right/left wrist, left/right leg, chest, and waist) and the angular velocity from two IMUs (chest and waist) [22]. Moreover, Mazilu et al. tested eleven machine learning models (e.g., random forests, k-nearest neighbor, and AdaBoost) on seven hand-engineered acceleration features (i.e., mean, standard deviation, variance, entropy, energy, FI, and power) [23]. Additionally, Shi et al. combined all the aforementioned features [21], [24]–[26] with wavelet features to form a set of 67 expressive features to characterize FOG [16]. They compared seven popular machine learning algorithms (e.g., k-nearest neighbors, support vector machines, and extreme gradient boosting (XGBoost)) and concluded that XGBoost enabled the best FOG detection performance [16].

However, manually engineered features run the risk of not being fully generalizable to all patients, given that PD and FOG are highly heterogeneous. Recent studies have thus shifted towards end-to-end DL models [14]–[16]. Due to their large parametric space, DL techniques can directly infer relevant features from raw input data. For example, Zhang et al. used raw acceleration and spectrograms of one waist IMU as input for a DeepCNN-LSTM model trained to detect FOG [27]. Li et al. proposed a DL model using a temporal convolutional network (TCN) and long-short-term-memory network for FOG detection using acceleration signals from three IMU sensors [28]. O’Day et al. fed raw acceleration and angular velocity data from one to eleven IMUs into a convolutional neural network (CNN) to detect FOG [15]. Lastly, Shi et al., besides proposing the feature-based model, also introduced an improved CNN method that used the continuous wavelet transform (CWT) as a pre-processing step on each acceleration and angular velocity signal to generate scalograms which were used as input for a CNN [16]. Their results showed that CWT, in combination with a CNN, is state-of-the-art in FOG detection. However, as illustrated in figure 1, all these prior studies that applied a window-based method may not be the most optimal for defining the exact onsets and offsets of FOG episodes and for differentiating between FOG manifestations that may both be present within the same time window.

**Fig. 1.**
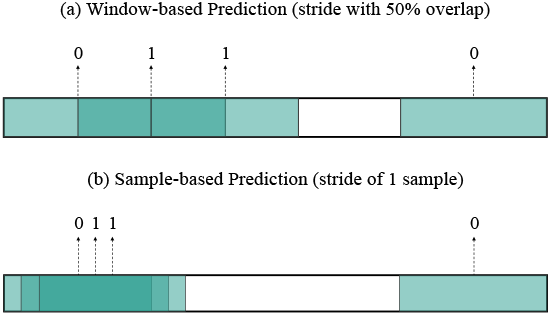
This example shows the difference between window- and sample-wise predictions for window-based models. The sliding windows were shown in green, with gradients representing the overlap. The x-axis represents the timeline for the annotations. This example shows that generating window-wise prediction with a 50% overlap between consecutive windows results in a downsampled prediction.

### B. Sample-based methods

Sample-based methods treat FOG detection as an action segmentation task [9], [29], [30]. These approaches distinguish between FOG and non-FOG on the sample level by generating one output for each input sample. Such sample-to-sample prediction eliminates the need for pre-defined window sizes and majority voting, which allows for more fine-grained activity detection [31]. In a recent study, we introduced the multi-stage temporal convolutional network combined with a many-to-one training scheme (MS-TCN) [9]. This temporal convolutional neural network architecture modified the training procedure of the multi-stage temporal convolutional network [32], initially proposed for video action segmentation, to improve FOG detection performance.

## III. Methods

In this study, we modified our MS-TCN model [9] to the FOG manifestation detection task. To evaluate the performance of our new approach, we compared it with three state-of-the-art window-based methods: feature-based [16], signal-based [15], and CWT-based [16] by extending each them to the FOG manifestation detection task. In the following sections we will first explain the gait tasks performed and the problem of FOG manifestation detection and its requirements. We will then discuss the implementation details of our proposed model, followed by an overview of the characteristics and implementation details of the three window-based models we used for comparison.

### A. Problem Definition

An IMU trial can be represented as *X ∈* ℝ^*T* ×*C*^*in*, where *T* is the number of samples, and *C*_*in*_ is the input feature dimension. Each IMU trial *X* is associated with a ground truth label vector *Y* ^*T* × *L*^, where the label *L* represents the manual annotation of FOG by the clinical experts. To generate predictions for each sample, sample-based methods learn a function 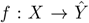 that transforms a given input sequence *X* = *x*_0_, …, *x*_*T −*1_ into an output sequence 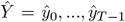 that closely resembles the manual annotations *Y*. Window-based methods split each IMU trial from *X ∈* ℝ^*T ×C*^*in* into multiple windows with a fixed number of samples equal to the window size *k* and generate a predicted label for each window, with the ground truth label for each window typically considered as the majority label within each window [15], [16]. A window-based model learns a function 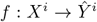 that transforms a given input sequence 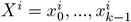 into an output label 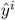 that closely resembles the ground truth label for window *i*.

### B. Sample-based method

Our model is an MS-TCN architecture [32] with two blocks that take a sequence of IMU signals as input and transforms them through multiple temporal convolutional layers. The first block is an initial prediction generation block that generates probabilities for each output label of a given sample. The second block is a prediction refinement block that contains multiple stages, each with multiple temporal convolutional layers, to refine the initial predictions and prevent oversegmentation errors [32]. Although MS-TCN enabled state-of-the-art activity recognition in various applications that deal with IMU data [33], [34], previous studies have shown that a many-to-one training strategy [35] enables improved generalization [35], [36]. Therefore, we train the prediction generation block with a many-to-one training scheme for our FOG detection model [9].

In the many-to-one training scheme, given a receptive field size *n*, each input IMU sequence is first replication-padded on both sides with (*n −* 1)*/*2, resulting in a sequence of length *T* + *n−* 1. The IMU sequence is then split into *T* chunks, each with size *n*, using a stride of one sample. These chunks are used to train the prediction generation block. The first layer is a 1 × 1 convolution layer that adjusts the input dimension from *n* × *C*_*in*_ into *n* × *C*, where *C* is the number of filters. The adjusted feature map is passed through four TCN blocks, each containing a dilated temporal convolutional layer [37], [38], batch normalization layer (BN) [39], ReLU activation function, and a residual connection [40]. These TCN blocks map the adjusted features to 1 × *C*. The output feature is passed through a 1 × 1 convolutional layer with a softmax activation function to output the initial probabilities for the *L* output classes. These initial probabilities are stacked together to form the initial prediction for each IMU sequence.

The initial probabilities of each IMU sequence of length *T* are fed into the prediction refinement block, which consists of *S* stages. Each stage refines the prediction from the previous stage using a series of TCN blocks. The first layer of each stage is a 1 × 1 convolution layer that adjusts the input dimension from *T × L* into *T* × *C*, where *C* is the number of filters. The adjusted features are passed through eight TCN blocks, each containing a dilated temporal convolution [37], [38], BN layer [39], ReLU function, and a residual connection. The last layer of each stage is a 1 × 1 convolutional layer with a softmax activation function to output refined probabilities for the *L* classes for each sample in time.

The same training procedure and model hyperparameters were used as in the original study [9].

### C. Window-based methods

#### 1) Feature-based Model

This study used the feature-based model proposed by Shi et al. [16], which applied the XG-Boost [41] algorithm on sixty-seven features generated from the IMU on the left tibia, including five frequency domain features, six entropy features, and 54 wavelet features. Two features calculated from magnetometer signals were removed as our dataset does not include magnetometers. The features were computed by following the same pre-processing procedure as the original study. Specifically, the accelerometer signals were filtered with a 4th-order Butterworth band-pass filter (0.2-15 Hz), and the angular velocity signals were filtered with a 4th-order Butterworth low-pass filter (10 Hz), at a sampling frequency of 50Hz. The window size was set to one second with 50% overlap between consecutive windows [16]. Instead of using majority voting to determine the ground-truth label, the centered label of each window was used as the ground truth to avoid changing the experts’ annotation [16]. The same training procedure and hyperparameters of the XGBoost model were used as in the original study.

#### 2) Signal-based Model

In addition to the feature-based model, we also used the signal-based model proposed by O’Day et al. [15]. The same pre-processing procedure was used as in the original study. Specifically, the IMU data was split into windows of two seconds with 50% overlap between consecutive windows. Each window was normalized to zero mean and unit variance and augmented with random rotations about the individual IMU axes to simulate variation in sensor placement [15]. The centered label of each window was used as the ground truth of that window. The same training procedure and hyperparameters of the CNN model were used as in the original study.

#### 3) CWT-based Model

Lastly, this study used the CWT-based model proposed by Shi et al. [16]. The same preprocessing procedure was used as in the original study. Specifically, the raw IMU signals were first normalized and split into multiple windows with a window size of four seconds and 50% overlap between consecutive windows. The normalized signals in each window were used to generate scalograms with CWT. The centered label of each window was used as the ground truth of that window. The same training procedure and hyperparameters of the CNN model were used as in the original study.

## IV. Evaluation

### A. Dataset

An existing IMU dataset was used [9]. The dataset includes twelve PD patients, all recruited if they subjectively reported having at least one FOG episode per day with a minimum duration of five seconds. Subjects varied in their age (mean: 69.33 years, range: 57–76), disease duration (mean: 12.33 years, range: 3–23), and self-reported FOG severity with New Freezing of Gait Questionnaire [42] (mean: 20.54, range: 12–26).

The dataset [9] was recorded with five Shimmer3 IMU sensors on all subjects, attached to the pelvis, both sides of the tibia and talus. All IMUs recorded at a sampling frequency of 64 Hz during the measurements. Synchronously, RGB videos were captured at 30 frames per second for offline FOG annotation purposes. FOG events were visually annotated at a frame-based resolution by a clinical expert, after which all FOG events were verified by another clinical expert using Elan annotation software [11]. Annotators used the definition of FOG as a brief episode with the inability to produce effective steps, and the episode ended at the foot off that was followed by at least two effective steps [1], [11], which adopts a stricter definition of FOG that distinguishes shuffling and festination as non-FOG events, and only trembling in place and complete akinesia as FOG events. The definition of shuffling was based on [8], namely small steps with minimal forward progression, while festination was defined as a tendency to move forward with increasingly rapid but ever smaller steps [2].

The dataset featured the timed up-and-go (TUG) test, with turning in both directions, and the 360 turning in-place (360Turn) test [43], with alternating 360-degree turning for one minute. The tasks were measured with and without a dual task, namely the auditory Stroop task [43], [44], and with and without a self-generated or researcher-imposed stopping. Stopping in the TUG was performed four times, twice with a stop in the straight walking part and twice with a stop in the turning part of the TUG; while stopping in the 360Turn was performed once. All pre-mentioned tasks were done first in the clinical off-medication state (approximately 12 hours after the last PD medication intake) and repeated in the same order during the on-medication state (approximately one hour after medication intake).

### B. Experimental Setting

The window-based models have several drawbacks. Firstly, they depend on majority voting, which alters expert annotations and affects FOG severity outcome values, as illustrated in figure 2a. Secondly, window-based models lack the granularity of expert observation to accurately identify the start and end of each FOG episode. Lastly, the window-based models were trained and evaluated without padding on both sides. As a result, these models would generate a prediction shorter than the original input sequence. As shown in figure 2b, even when generating sample-wise predictions by sliding with one sample, they would still predict a shorter sequence of length *T − k* + 1 given an input sequence of length *T* and window size of *k*.

**Fig. 2.**
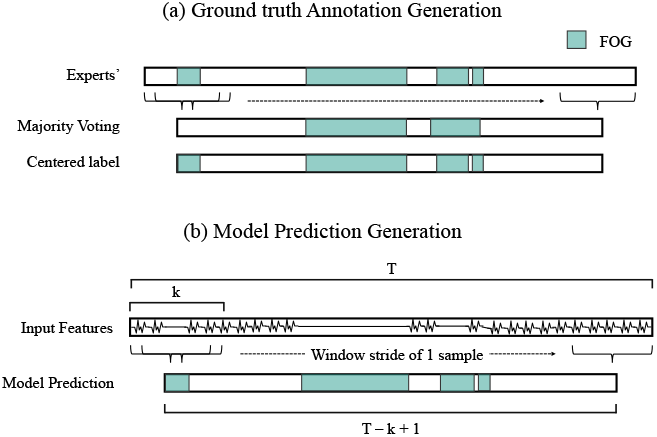
Visual representation highlighting how window-based FOG detection methods alter the ground-truth experts’ annotation. Figure (a) shows that majority voting results in minor temporal shifts segments and the removal of short segments. In contrast, using each window’s centered label as ground truth maintains the experts’ annotation. (b) Shows that shifting each prediction window with a stride of 1 sample enables fine-grained sample-wise predictions, but still reduces the sequence from length *T* to *T − k* + 1, given a window of duration *k*.

To overcome these issues, we defined a uniform evaluation setting for comparing the models. Firstly, we addressed the issue of adapting the experts’ annotations by defining the ground truth label as the center label of the window. This consistent ground truth labeling approach was employed during model training and inference, ensuring coherence and comparability. Secondly, to achieve consistent granularity in predictions, we employed a sliding window technique with a stride of 1 sample for window-based methods during inference. Meanwhile, during model training, we maintained a 50% overlap approach. This methodology ensured that predictions were generated at the same granularity as our sample-based model, enhancing the evaluation consistency. Thirdly, when comparing the performance of MS-TCN with other models, we evaluated the *T − k* + 1 predicted sequence. This approach was adopted to avoid evaluating models with varying lengths of ground truth sequences, as such differences can introduce discrepancies, particularly when the original *T* sequence contains sitting events that may not be present in the shortened sequence. Conversely, while further investigating the performance of MS-TCN in discerning FOG manifestations from other forms of volitional movement cessation, the entire *T* predicted sequence was evaluated. Fourthly, each method was evaluated for multiple different window durations for window-based methods and receptive fields for sample-based methods. This comprehensive evaluation accounted for varying temporal contexts and allowed a more thorough analysis of the model’s performance. Lastly, except for the feature-based model, all models were trained using data from all five IMUs. The feature-based model exclusively employed features derived from the left tibia IMU (denoted as “leg” in [16]).

All 346 trials in the dataset were used to train and evaluate the models. The labels for the FOG manifestation detection task were three (*L* = 3), with *l* = 0 for non-FOG (i.e., walking, sit-to-stand, stand-to-sit, and other volitional movement cessations), *l* = 1 for trembling in place, and *l* = 2 for complete akinesia. MS-TCN was additionally evaluated based on its performance in discerning FOG manifestations from other types of movement cessation, such as volitional stopping and sitting. For this task, MS-TCN was trained with five target classes (*L* = 5), where *l* = 1 represents trembling in place, *l* = 2 represents complete akinesia, *l* = 3 represents stopping, *l* = 4 represents sitting, and *l* = 0 represents all other events (i.e., walking, sit-to-stand, and stand-to-sit). All other events are hereinafter simply referred to as “walking”. All experiments were conducted by following a leave-one-subject-out cross-validation approach. Specifically, the dataset was partitioned into training and testing sets, where one subject served as the testing set and the remaining subjects as the training set. This procedure was repeated iteratively until each subject had been evaluated.

### C. Metrics

This paper assessed FOG severity from a clinical perspective, primarily focusing on two outcomes: percentage time-frozen (%TF) and the number of detected FOG episodes (#FOG) [10]. To further quantify the FOG manifestations, this study proposed the percentage time of trembling in place (%TF-T) and percentage time of complete akinesia (%TF-A), inspired by previous studies [45], [46]. The (%TF-T) was calculated as the total duration of trembling in place divided by the total duration of all tasks. The %TF-A was calculated as the total duration of complete akinesia divided by the total duration of all tasks. Table I summarizes the FOG severity for each subject in the dataset. To assess the agreement between model-predicted FOG severity and expert-annotated FOG severity, the intra-class correlation coefficient (ICC(2,1)) was used. The ICC value indicates the agreement between the model and the experts. A higher ICC value suggests higher agreement. According to [47], the agreement strength was classified as follows: *≥*0.80: strong, 0.6-0.79: moderately strong, 0.3-0.59: fair, and *<*0.3: poor. As the clinical metrics are a summary of FOG severity per subject and insufficiently sensitive for model comparison [9], the F1 score was used to compare the performance of the different models.

**TABLE I.** DATASET CHARACTERISTICS

|  | #Trials | Duration | #FOG-Trials | #Trembling-Trials | #Akinetic-Trials | %TF | %TF-T | %TF-A | #FOG | #FOG-T | #FOG-A |
| --- | --- | --- | --- | --- | --- | --- | --- | --- | --- | --- | --- |
| S1 | 29 | 17.10 | 16 | 8 | 15 | 19.56 | 1.50 | 18.06 | 35 | 10 | 26 |
| S2 | 29 | 13.90 | 9 | 7 | 8 | 12.64 | 5.64 | 7.00 | 34 | 16 | 18 |
| S3 | 31 | 13.22 | 6 | 5 | 1 | 7.10 | 6.93 | 0.17 | 37 | 36 | 1 |
| S4 | 27 | 10.48 | 12 | 12 | 0 | 7.89 | 7.89 | 0.00 | 30 | 30 | 0 |
| S5 | 16 | 6.88 | 4 | 4 | 1 | 9.80 | 8.73 | 1.07 | 22 | 22 | 1 |
| S6 | 32 | 12.84 | 1 | 1 | 0 | 0.10 | 0.10 | 0.00 | 1 | 1 | 0 |
| S7 | 32 | 17.64 | 22 | 22 | 2 | 14.10 | 13.82 | 0.27 | 106 | 104 | 2 |
| S8 | 33 | 13.48 | 0 | 0 | 0 | 0.00 | 0.00 | 0.00 | 0 | 0 | 0 |
| S9 | 31 | 14.51 | 15 | 15 | 0 | 4.49 | 4.49 | 0.00 | 61 | 61 | 0 |
| S10 | 21 | 25.59 | 21 | 20 | 13 | 52.86 | 35.53 | 17.32 | 111 | 103 | 25 |
| S11 | 31 | 15.61 | 17 | 17 | 5 | 12.27 | 10.65 | 1.62 | 74 | 66 | 8 |
| S12 | 34 | 20.40 | 10 | 6 | 7 | 2.07 | 0.79 | 1.28 | 19 | 9 | 10 |
| Overall | 346 | 181.70 | 133 | 117 | 52 | 14.62 | 9.58 | 5.04 | 530 | 458 | 91 |

Overview of the dataset for each subject, including the number of trials, total duration in minutes, the number of FOG trials (#FOG-trials), the number of trembling trials (#Trembling-Trials), the number of akinetic trials (#Akinetic-Trials), the percentage of time frozen (%TF), the percentage of time trembling (%TF-T), the percentage of time akinetic (%TF-A), the number of FOG episodes (#FOG), the number of trembling episodes (#FOG-T), and the number of akinetic episodes (#FOG-A). The #FOGs are not the sum of #FOG-T, and #FOG-A, as a FOG episode could contain both manifestations.

The F1 score is a widely used metric for evaluating the accuracy of binary classification models. For sample-wise predictions, the comparison is performed at the individual sample level. Each prediction of the sample is classified as True Positive (TP), False Positive (FP), or False Negative (FN) based on the correspondence between the predicted and ground truth labels. The F1 score is calculated under the formula: 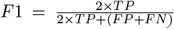. For the tasks of multi-class manifestation classification (normal movement, trembling in place, and complete akinesia) and multi-class manifestation and volitional movement cessation classification (normal movement, trembling in place, complete akinesia, stopping, and sitting), we calculated an F1 score for each class individually in a one vs. all manner. This means that when computing the F1 score for a specific class, that class is considered positive, while all other classes are treated as negative. These individual F1 scores were then averaged (F1-Total) for each subject [48].

### D. Statistics

The repeated measures anova test [49] was used to investigate whether the differences between the models in the F1 scores were statistically significant. When a significant difference was found, post hoc paired Student’s t-tests [50] were applied to investigate significant differences between pair-wise models. The post hoc hypotheses were corrected for multiple comparisons, as defined in Li [51]. The homogeneity of variances was verified in all metrics across subjects with Levene’s tests [52]. The Shapiro-Wilk test [53] was used to determine whether the variables were normally distributed across subjects. The Bland–Altman plot [54] was used to investigate systematic bias between FOG severity outcomes (i.e., %TF, #FOG, %TF-T, and %TF-A) predicted by MS-TCN and the experts’ annotation. The significance level for all tests was set at 0.05. All analyses were performed using SciPy 1.7.11, bioinfokit 2.1.0, statsmodels 0.13.2, and pingouin 0.3.12, written in Python version 3.7.11. The post hoc test was performed using scmamp 0.2.55 [55] written in R version 4.0.3.

## V. Results

### A. FOG manifestation detection: comparison with three baseline models

To compare the performance of the MS-TCN model with the three baseline models in detecting FOG manifestations, we trained and evaluated the models with three different window/receptive field sizes (i.e., one, two, and four seconds). Table II displays the F1-Total, F1-Trembling, and F1-Akinetic for the four models. The results indicated that the feature-based, signal-based, and CWT-based models trained with a four-second window size and MS-TCN trained with a four-second receptive field size achieved the highest F1-Total and F1-Trembling. The FOG manifestation detection performance of these four best-performing models was statistically compared, which showed statistical significance for the F1-Total (*p* = 0.0002). Figure 3 presents the box plot of the results, showing that MS-TCN outperformed the other three models in terms of the F1-Total. Moreover, the post hoc tests confirmed that the difference between MS-TCN and the other three models in terms of F1-Total was statistically significant.

**TABLE II.** COMPARISON OF THE FOUR MODELS IN TERMS OF THE F1 SCORE

| Model | Size (s) | F1-Trembling | F1-Akinetic | F1-Total |
| --- | --- | --- | --- | --- |
| Feature-based [16] | 1 | 0.51 | **0.82** | 0.66 |
|  | 2 | 0.54 | 0.81 | 0.68 |
|  | 4 | **0.56** | 0.81 | **0.69** |
| Signal-based [15] | 1 | 0.23 | 0.21 | 0.22 |
|  | 2 | 0.29 | 0.27 | 0.28 |
|  | 4 | **0.33** | **0.36** | **0.34** |
| CWT-based [16] | 1 | 0.56 | **0.79** | 0.68 |
|  | 2 | 0.58 | 0.58 | 0.58 |
|  | 4 | **0.63** | 0.77 | **0.70** |
| MS-TCN [9] | 1 | 0.72 | **0.83** | 0.77 |
|  | 2 | 0.73 | 0.82 | **0.78** |
|  | 4 | **0.74** | 0.82 | **0.78** |

We compared the proposed MS-TCN model with three window-based models to detect two FOG manifestations. The $T - k + 1$ predicted sequences of all models were compared with the ground truth annotation. The largest $k$ was set for all models to maintain fair comparison, i.e., $k = 4$ .

**Fig. 3.**
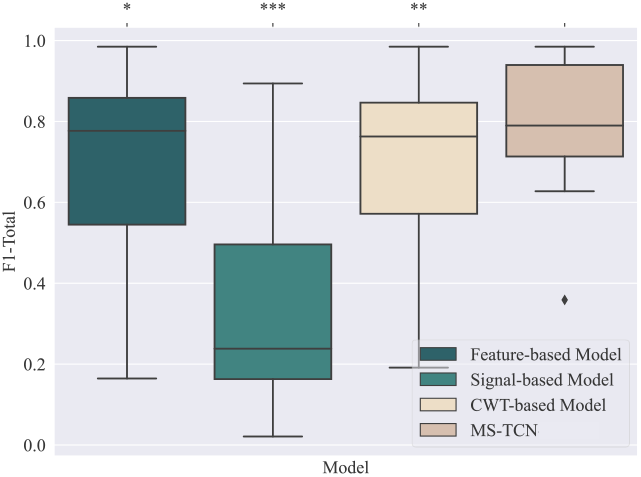
The spread of the F1-Score across subjects. The anova test showed a significant difference between the F1-Score metrics of the four models(p=0.0002). The significance levels of the post hoc tests with respect to the MS-TCN model (corrected for three pairwise comparisons are visualized above their respective boxplot. Significance levels were visualized as: p*≤*0.005 (***), p*≤*0.01 (**), and p*≤*0.05 (*).

Additionally, as shown in Figure 4, both the signalbased and feature-based models had many instances of oversegmentation. While the CWT-based model had fewer errors, it incorrectly classified akinetic FOG episodes as trembling and did not detect short trembling FOG episodes. This shows that the MS-TCN model not only outperforms these models in terms of the F1 scores but that it is also able to predict FOG manifestations with fewer over-segmentation errors.

**Fig. 4.**
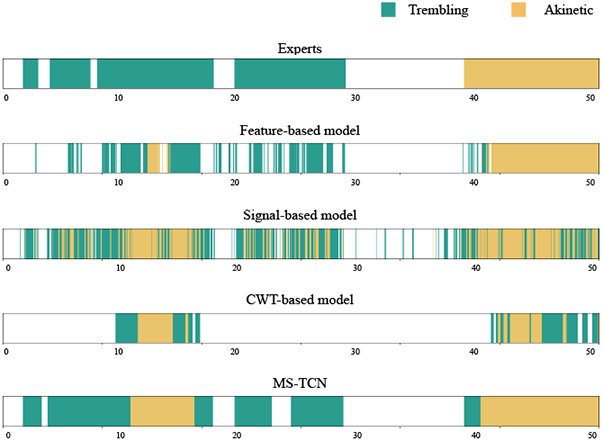
Overview of the predictions of the best four models compared with the experts’ annotation. The figures visualize the over-segmentation of the window-based models. The x-axis denotes the time of the trial in seconds.

### B. FOG manifestation severity assessment

Next, we evaluated the MS-TCN in terms of FOG manifestation severity outcomes. Results showed a strong agreement between the model and experts in terms of %TF (ICC = 0.96, CI=[0.79,0.99]), #FOG (ICC = 0.84, CI=[0.55,0.95]), and %TF-A (ICC = 0.87, CI=[0.63,0.96]), and a moderately strong agreement in terms of %TF-T (ICC = 0.64, CI=[0.16,0.88]). The Bland–Altman plots presented in figure 5 demonstrated that our model systematically underestimated the %TF (2.42% (CI=[0.44,4.40])) and showed no systematic error for the other three clinical metrics, with a mean bias of 5.83 (CI=[*−*6.65,18.32]) for #FOG, 3.01% (CI=[*−*0.77,6.78]) for %TF-T, and *−*0.59% (CI=[*−*3.08,1.90]) for %TF-A. Notably, %TF was underestimated mainly due to two subjects using walking aids, as there was no systematic error when evaluating only the remaining subjects. Moreover, the limits of agreement (LOA) of %TF-T were *−*8.63% (CI=*−*15.17,*−*2.10) to 14.65% (CI=[8.11,21.19]), and for %TF-A the LOA were *−*8.27% (CI=[*−*12.58,*−*3.95]) to 7.09% (CI=[2.78,11.40]). The agreement was lower for trembling than for akinetic FOG with broader LOA, showing that the model made more errors in detecting trembling in place than complete akinesia.

**Fig. 5.**
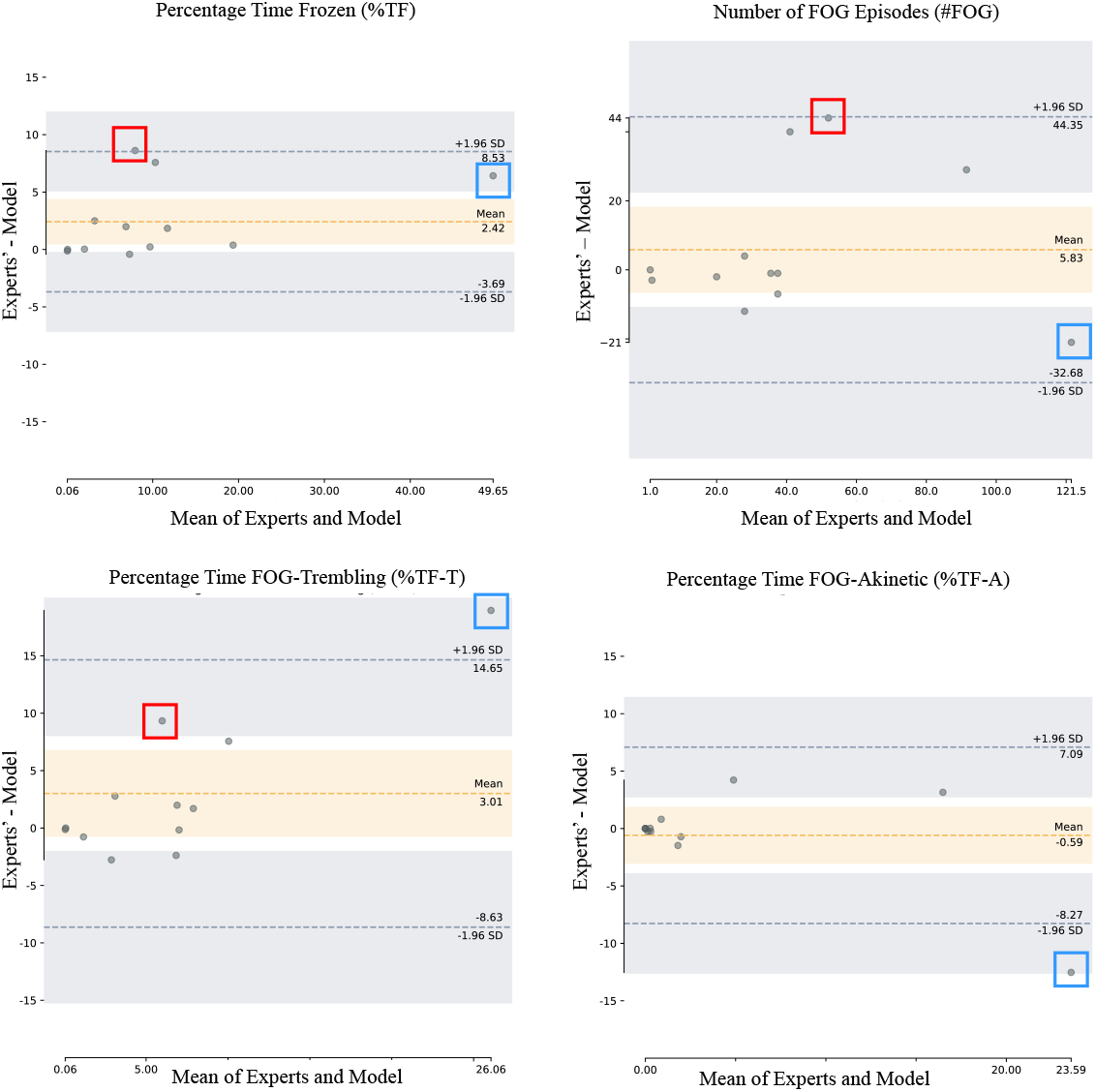
The Bland-Altman plot compares the scores of four clinical metrics from MS-TCN and experts. The dots represent the difference in scores per patient on the y-axis (i.e., model’s %TF, #FOG, %TF-T, or %TF-A subtracted from experts’ %TF, #FOG, %TF-T, or %TF-A), plotted against the mean score per patient from the model and the experts on the x-axis. The orange area shows the 95% CI for the mean bias, while the gray area shows the 95% CI for the upper and lower limits of agreement. No systematic error was found in #FOG, %TF-T, and %TF-A, while a systematic error was found in %TF. Two subjects using mobility aids are indicated with colored blocks (S10: blue; S11: red). For S10, who has a high #FOG, the model predicted a lot of trembling episodes as akinetic. Whereas for S11, the model predicted a lot of short trembling episodes as non-FOG.

### C. FOG manifestations versus other forms of volitional movement cessation

Next, we investigated the proposed model’s ability to distinguish FOG manifestations from volitional movement cessation by training the model with five target classes, i.e., walking, trembling, akinetic, stopping, and sitting. As seen in the confusion matrix (Figure 6), the model correctly predicted 94% of the walking samples, 71% of the akinetic samples, 63% of the stopping samples, and 86% of the sitting samples. However, the model struggled to accurately identify trembling samples, with only 42% of them correctly classified, while 29% were classified as walking and 21% as akinetic. To investigate the model’s ability to distinguish between stopping and FOG manifestations, we split up the results for non-FOG and FOG trials. When evaluating non-FOG trials, the model could accurately annotate 80% of stopping samples as stopping. In contrast, when evaluating FOG trials, the model can only correctly annotate 42% of stopping samples, with 24% as non-FOG, 14% as trembling, and 18% as akinetic. These results demonstrate that the model can accurately detect stopping in non-FOG trials but had difficulty in trials that contained FOG segments. These phenomena are demonstrated in the qualitative results presented in Figure 7. Observe in figure 7c that the model made errors distinguishing between trembling and akinetic in trials where both manifestations were present and in figure 7f between akinetic and stopping in trials where both appeared.

**Fig. 6.**
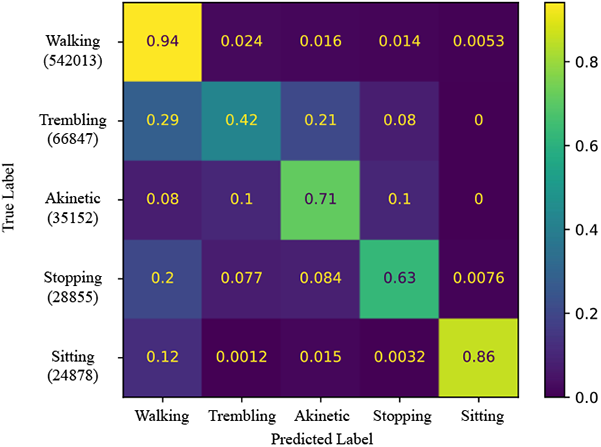
The normalized confusion matrix to visualize the ability of the MS-TCN in distinguishing the five classes. The confusion matrix shows the number of TP, TN, FP, and FN for each class. The true label of each sample (with the amount of true labels) is shown at the start of each row, and the predicted label is shown at the bottom of each column. We normalized the confusion matrix by dividing each element by the total number of samples in the corresponding true class to show the ratio of correct predictions for each class.

**Fig. 7.**
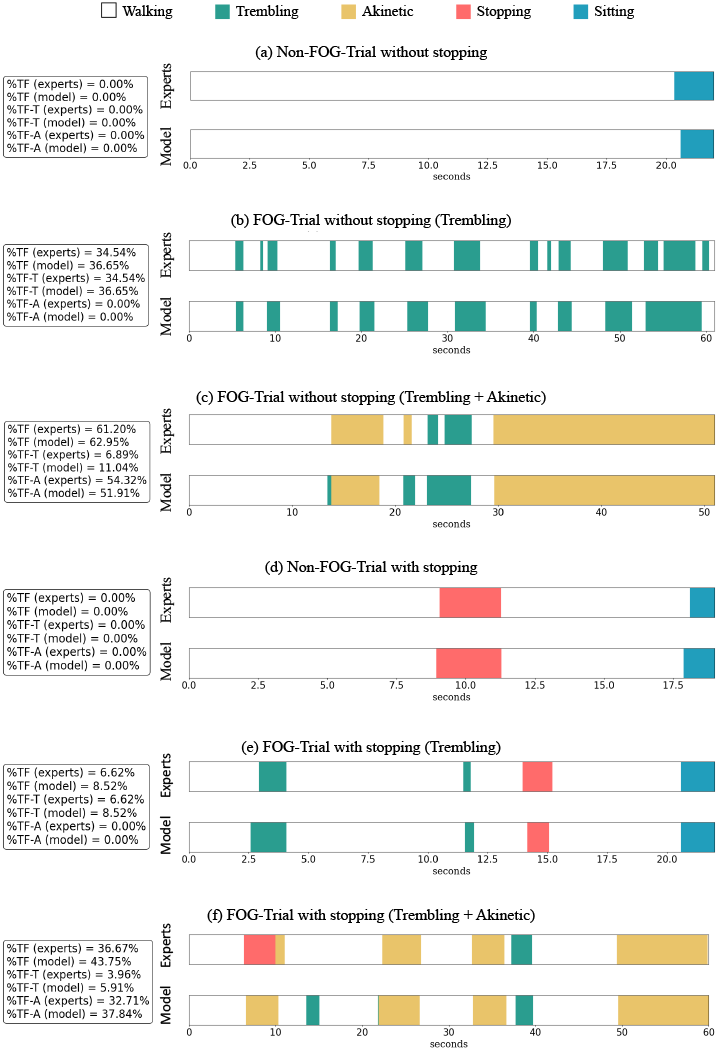
Overview of the annotations for six IMU trials. Six trials include six different types of experts annotations: a) Non-FOG trial without stopping, b) FOG trial without stopping (only Trembling), c) FOG trial without stopping (Trembling + Akinetic), d) Non-FOG trial with stopping, e) FOG trial with stopping (only Trembling), and f) FOG trial with stopping (Trembling + Akinetic). The figures visualize the difference between the manual segmentation by the experts (top) and the automated segmentation by the MS-TCN model (bottom), with white=walking, green=trembling, yellow=akinetic, red=stopping, and blue=sitting. The x-axis denotes the time of the trial in seconds.

## VI. Discussion

Previous FOG assessment studies [9], [15], [16], [29] combined various types of FOG into a single category. However, FOG can have different manifestations, which may have other pathophysiologic origins [8]. Therefore, objectively detecting these different FOG manifestations is crucial to tailor future FOG treatment approaches. To address this bottleneck, this study extended the state-of-the-art MS-TCN model [9] to support the detection of two FOG manifestations, i.e., trembling and akinetic FOG. Our proposed model was quantitatively compared to three state-of-the-art window-based models [15], [16] by extending these models to support manifestation detection. Results showed that our model statistically outperforms these models on FOG manifestation detection in terms of the total F1 score. Notably, the window-based models we utilized were not explicitly trained to minimize over-segmentation errors; hence, we did not evaluate them using the Segmentwise F1 score [38], which effectively penalizes such errors. Nevertheless, through a qualitative analysis of the predicted annotations, we observed that our model exhibits fewer oversegmentation errors when predicting FOG manifestations.

To quantify the severity of FOG manifestations, previous studies calculated the percentage of each FOG manifestation with respect to the total duration of FOG [45] and the number of episodes of each manifestation separately [46]. Nevertheless, when different experts annotate the same percentage of FOG manifestation but with varying total FOG durations, it can result in different durations for each manifestation. This implies that using the percentage of each manifestation within observed FOG as a metric to quantify the severity of FOG manifestations may not be reliable. As a result, inspired by previous studies [45], [46], we proposed two metrics, i.e., %TF-T and %TF-A, to quantify FOG manifestation severity. Our proposed model showed a strong agreement with the experts’ observations for %TF-A (ICC=0.87) and a moderately strong agreement for %TF-T (ICC=0.64). The ICC for FOG manifestation severity between independent raters was reported as 0.31 (CI=[0.11,0.49]) for the percentage of trembling and 0.44 (CI=[0.35,0.54]) for the percentage of akinetic [45]. Although [45] showed that annotating FOG manifestations are challenging, which would result in a low inter-rater agreement, our model prediction showed a moderate to strong agreement with our experts’ annotation, showing its ability to learn how our experts’ annotated the trials.

Next, we investigated the performance of our model in distinguishing FOG manifestations from other forms of movement cessation, i.e., volitional stopping and sitting, by evaluating the model trained explicitly for the five classes: walking, trembling, akinetic, stopping, and sitting. Results showed that our model could correctly detect sitting from FOG manifestations. However, stopping could only be accurately detected in trials that do not contain FOG. More specifically, the model made more errors distinguishing between akinetic, trembling, and stopping in trials where all classes appeared. Hence, motor signals alone may be insufficient to distinguish stopping from FOG, particularly during complex motion sequences that are likely to be encountered in everyday life. A promising avenue is to amalgamate motor and physiological signals (e.g., heart rate), which have recently shown potential in distinguishing between FOG and stopping, but lack the expressivity to distinguish between FOG and gait [12], which was highly distinguishable in our approach. Therefore, including physiological signals in our method seems a promising future improvement.

Furthermore, the results showed that the agreement between our model in terms of trembling was lower than the agreement for akinetic. This finding aligns with previously reported lower inter-rater ICC values for trembling compared to akinetic FOG [45]. Trembling FOG (i.e., alternating tremulous oscillations with no forward progression) and akinetic FOG (i.e., no visible movement in the lower limbs) are determined based on observable leg motion. There are several potential explanations: Firstly, some trembling movements may not be observable in the videos by the experts, especially if the movements are very small. Although our study procedure had participants wearing tight-fitting shorts, this may become even more challenging in clinical practice where patients with FOG are wearing their own comfortable long-legged pants. Secondly, as FOG manifestations may shift within one episode, it becomes very challenging and time-consuming for the experts to label it to the highest detail. Therefore, they resort to labeling the episode (or larger blocks of the episode) to the manifestation that is dominantly present.

Several limitations in this study should be considered.

Firstly, the dataset used in this study consisted of videos annotated sequentially by two clinical experts, with the second expert verifying and correcting the annotations made by the first expert. Due to our sequential annotation process, there was no opportunity to measure inter-rater agreement in terms of %TF-T and %TF-A to compare against our models’ annotations. The second limitation is the limited amount of FOG manifestations present in the dataset. Specifically, the dataset contained only 17.41 minutes of trembling episodes and 9.16 minutes of akinetic episodes, within a total dataset duration of 181.7 minutes. Given that FOG occurrences are generally less frequent than non-FOG instances during inlab measurements [43], and akinetic FOG tends to occur less frequently compared to trembling [8], [45], the ratio of the two manifestations in our dataset was considered reasonable. However, future studies could explore larger datasets with more FOG samples, especially for complete akinesia, to provide more training data for DL models.

## VII. Conclusion

The current study is the first attempt to automatically quantify FOG manifestations using DL. Our approach outperforms three state-of-the-art FOG detection models and demonstrated a strong agreement with experts’ annotations on %TF, #FOG, and %TF-A and a moderately strong agreement for %TF-T. Future work is now possible to establish whether these results hold for a larger and more varied verification cohort.

## Data Availability

Our neural network architecture was implemented in Python 3.7 with Pytorch version 1.8.0 by adopting the public code repositories of Farha and Gall [1], Pavllo et al. [2], and O'Day et al [3]. The datasets analyzed during the current study are not publicly available due to restrictions on sharing subject health information.

https://github.com/yabufarha/ms-tcn

https://github.com/facebookresearch/VideoPose3D

https://github.com/stanfordnmbl/imu-fog-detection/

## Acknowledgment

We thank the participants for their willingness to participate.

## Notes

Po-Kai Yang was supported by the Ministry of Education (KU Leuven–Taiwan) scholarship. Benjamin Filtjens was supported by KU Leuven Internal Funds Postdoctoral Mandate PDMT2/22/046.

### Competing Interest Statement

The authors have declared no competing interest.

### Funding Statement

This study is funded by the KU Leuven Industrial Research Fund. Po-Kai Yang was supported by the Ministry of Education (KU Leuven Taiwan Scholarship). Benjamin Filtjens was supported by KU Leuven Internal Funds Postdoctoral Mandate PDMT2/22/046.

### Author Declarations

Ethics committee Research UZ/KU Leuven approved this study (S65059).

